# Interplay of PD-L1, FOXP3, and CD8 in the Immune Microenvironment of Penile Squamous Cell Carcinoma: Expression Profiles and Correlations

**DOI:** 10.1101/2024.12.11.24318871

**Authors:** Sofia Canete-Portillo, Antonio L. Cubilla, George J. Netto, Alcides Chaux

**Affiliations:** The University of Alabama at Birmingham, Birmingham, AL; Instituto de Patología e Investigación, Asunción, Paraguay; Perelman School of Medicine at the University of Pennsylvania, Philadelphia, PA; Facultad de Medicina, Universidad del Norte, Asunción, Paraguay

**Keywords:** Penile cancer, PD-L1, FOXP3, CD8, tumor microenvironment, immunotherapy

## Abstract

**Background:** Penile squamous cell carcinoma (PSCC) presents significant challenges in diagnosis and treatment. Understanding the complex immunological landscape, particularly the interaction between PD-L1, FOXP3, and CD8, is crucial for developing effective therapeutic strategies.

**Methods:** We analyzed 108 PSCC cases through 528 tissue microarray samples. Immunohistochemical staining was performed using standardized protocols to detect PD-L1, FOXP3, and CD8 expression. Expression patterns were evaluated in both tumor and stromal compartments, with assessment of membranous staining for PD-L1 and quantification of FOXP3+ and CD8+ lymphocytes.

**Results:** PD-L1 expression in tumor cells showed a moderate positive correlation (r=0.477, P<0.001) with CD8+ T-cell infiltration in the tumor compartment, suggesting an adaptive immune resistance mechanism. PD-L1 expression in tumor cells demonstrated a weak positive correlation (r=0.289, P=0.002) with FOXP3 expression in tumor-associated lymphocytes. The strongest correlation observed (r=0.717, P<0.001) was between FOXP3 expression in stromal lymphocytes and CD8+ T-cell presence in stromal areas. PD-L1 expression in lymphocytes showed a significant correlation (r=0.354, P<0.001) with FOXP3 expression in stromal lymphocytes.

**Conclusions:** Our findings reveal complex interactions between PD-L1, FOXP3, and CD8 within the tumor microenvironment of PSCC. The strong correlation between stromal FOXP3+ and CD8+ cells points to a crucial immune regulatory axis that could influence treatment outcomes. These results provide insights for developing targeted therapeutic strategies and improving patient care in PSCC.

## INTRODUCTION

Penile squamous cell carcinoma (PSCC) represents a rare but aggressive malignancy that poses significant challenges in diagnosis, treatment, and management. Despite its relative rarity, PSCC carries substantial morbidity and mortality, particularly in developing countries where its prevalence is higher (Ahmed et al., 2020; Chaux et al., 2013). Understanding the complex immunological landscape of PSCC has emerged as a critical area of study, offering potential insights into disease progression, treatment response, and novel therapeutic approaches.

The tumor microenvironment (TME) in PSCC exhibits distinct characteristics compared to other squamous cell carcinomas, particularly regarding immune cell composition and marker expression (Müller et al., 2021). Three key components of the immune response have emerged as crucial players in this context: programmed death-ligand 1 (PD-L1), forkhead box P3 (FOXP3), and cluster of differentiation 8 (CD8). PD-L1 serves as a critical immune checkpoint molecule that tumors can exploit to evade immune detection. FOXP3 is a transcription factor predominantly expressed in regulatory T cells (Tregs), which are known to suppress antitumor immune responses and promote an immunosuppressive microenvironment. CD8 is a marker for cytotoxic T lymphocytes, which are essential effector cells in the antitumor immune response (Ottenhof et al., 2018).

Recent studies have demonstrated that PD-L1 expression in PSCC varies significantly, with positivity rates ranging from 40% to 62.2% (Warli et al., 2022). This variability may reflect different methodological approaches and scoring systems, but also suggests the complex nature of immune regulation in these tumors. The presence of FOXP3+ Tregs within the TME has been associated with immune suppression and poorer clinical outcomes, while higher levels of CD8+ T cell infiltration generally correlate with better prognoses (Yezhong Tang et al., 2022).

The relationship between these immune markers is further complicated by the presence of tertiary lymphoid structures (TLS), organized aggregates of immune cells that can influence local immune responses and potentially affect treatment outcomes (Tagami et al., 2022). Additionally, the impact of human papillomavirus (HPV) status on immune marker expression adds another layer of complexity to understanding the immune landscape of PSCC (Mumba, 2024).

The interplay between these immune components is critical in shaping the overall immune response to the tumor and potentially influencing disease progression and treatment outcomes (Deng et al., 2017). Understanding these complex interactions could provide valuable insights into the mechanisms of immune evasion in PSCC and potentially identify new targets for immunotherapy (Vries et al., 2023).

Our study aims to conduct a comprehensive examination of the expression profiles and correlations between PD-L1, FOXP3, and CD8 within both tumor and stromal compartments of PSCC. By analyzing these relationships, we seek to: (1) characterize the immune landscape of PSCC in terms of PD-L1 expression, regulatory T cell presence, and cytotoxic T cell infiltration; (2) identify potential patterns of immune marker expression that could serve as prognostic or predictive biomarkers; (3) explore the relationships between these immune markers to better understand the mechanisms of immune regulation in PSCC; and (4) provide a foundation for the development of targeted immunotherapeutic strategies in PSCC.

## MATERIALS AND METHODS

### Study Design and Ethical Considerations

The current study was approved by the Institutional Review Board at the Johns Hopkins School of Medicine (Baltimore, MD). All samples were obtained with appropriate ethical approvals and patient consents, and the study was conducted in accordance with the Declaration of Helsinki.

### Case Selection and Tissue Microarray Construction

The study analyzed tissue samples from 108 patients with invasive squamous cell carcinoma of the penis from the consultation files of one of the authors (ALC). Cases were selected based on availability of formalin-fixed, paraffin-embedded tissue blocks, with 1–4 blocks selected from each case to ensure adequate representation of tumor heterogeneity. Four tissue microarrays (TMA) were constructed at the Johns Hopkins TMA Lab Core (Baltimore, MD) using established procedures (Fedor & De Marzo, 2005). Three tissue cores of 1 mm each were obtained per block, providing 3–12 spots per case. Normal tissue from various anatomical sites were included as control tissue. A total of 528 TMA spots were evaluated from the 108 cases, ensuring robust sampling for statistical analysis.

### Morphological Evaluation

Pathologic features were evaluated using H&E-stained tissue sections by three expert genitourinary pathologists (ALC, GN, AC). Histologic subtyping was performed on whole tissue sections using the 5th edition (2022) WHO criteria for classification of tumors of the urinary system and male genital organs (Moch et al., 2022). Histologic grading was conducted spot by spot using previously published and validated criteria (Chaux, 2015). Grade 1 tumors showed well-differentiated cells with minimal basal/parabasal cell atypia. Grade 3 tumors demonstrated anaplastic cells with nuclear pleomorphism, coarse chromatin, prominent nucleoli, irregular and thickened nuclear membranes, and abundant atypical mitoses. Grade 2 tumors represented an intermediate category, not fitting criteria for grades 1 or 3. Host response was evaluated spot by spot, with inflammatory infiltrate intensity classified as none, mild, moderate, or intense (Cocks et al., 2017).

### Immunohistochemical Analysis

Immunohistochemical staining was performed using standardized protocols on an automated immunostainer to ensure consistency and reproducibility. PD-L1 antibody (Cell Signaling, E1L3N) was applied at 1:100 dilution with a 45-minute room temperature incubation. CD8 antibody (Thermo Scientific, RB-9009-P0) was used at 1:800 dilution with a 45-minute room temperature incubation, following EDTA buffer (pH 9.0) antigen retrieval. FOXP3 antibody (clone 236A/E7, Abcam) was applied following manufacturer’s protocols. For all antibodies, heat-induced antigen retrieval was performed for 20 minutes in a steamer with citrate buffer solution (pH 6.0). Detection utilized antimouse or antirabbit horseradish peroxidase–conjugated secondary antibodies with 3,3′-diaminobenzidine counterstaining.

### Expression Analysis and Scoring

Expression analysis was performed independently by two experienced pathologists (SCP, GN) blinded to clinical data. PD-L1 positivity was defined as any extent of tumor cell or immune cell positivity in representative TMA spots (>0%). CD8+ and FOXP3+ lymphocytes were quantified in both tumor and stromal compartments, evaluating the areas of highest density per high-power field (×40) for both intratumoral and peritumoral distributions. In cases of scoring discrepancy, consensus was reached through joint review.

### Statistical Methods

Statistical analyses were performed using Python 3.12.8 to evaluate correlations between different immune markers. Pearson correlation coefficients were calculated to assess relationships between marker expressions in various compartments. Specifically, correlations were analyzed between PD-L1 expression in tumor cells and CD8+ T-cell infiltration, PD-L1 expression and FOXP3+ lymphocytes, and FOXP3+ stromal lymphocytes and CD8+ T-cells in stromal areas. P-values less than 0.05 were considered statistically significant. Additional analyses included assessment of marker expression patterns in relation to histological grades and host response categories.

## RESULTS

### Patient and Tumor Characteristics

Our study analyzed 108 cases of invasive penile squamous cell carcinoma through tissue microarray analysis comprising 528 spots. The tumors represented various histological subtypes including Usual (predominant), Warty, Basaloid, and Warty-Basaloid types. Histological grades ranged from Grade 1 to Grade 3, with Grade 2 and 3 being most frequent.

### PD-L1 Expression Patterns

PD-L1 expression demonstrated marked heterogeneity in both tumor cells and tumor-infiltrating lymphocytes. In tumor cells, PD-L1 showed predominantly cytoplasmic/membranous staining, with expression levels ranging from 0% to 100%. Several cases exhibited high PD-L1 expression (>90%), particularly in Usual-type carcinomas. PD-L1 expression in lymphocytes ranged from 0 to 70%, with most cases showing values between 0-30%. Notably, PD-L1 expression in tumor cells demonstrated a moderate positive correlation with CD8+ T-cell infiltration (r=0.477, P<0.001), suggesting an adaptive immune resistance mechanism. This finding aligns with previous observations in other squamous cell carcinomas (Müller et al., 2021) and supports the concept of immune checkpoint activation in response to cytotoxic T-cell presence.

### FOXP3 Expression and Distribution

FOXP3+ cell distribution showed distinct patterns between tumor-associated and stromal compartments. Stromal FOXP3+ lymphocyte counts varied widely, ranging from 0 to more than 140 cells per high-power field, with most cases demonstrating strong staining intensity. Tumor-associated FOXP3+ lymphocytes were generally less numerous, ranging from 0 to approximately 56 cells per high-power field, indicating compartment-specific regulation of regulatory T-cell infiltration. FOXP3 expression in tumor-associated lymphocytes showed a weak positive correlation with PD-L1 expression in tumor cells (r=0.289, P=0.002). More significantly, FOXP3 expression in stromal lymphocytes exhibited a strong positive correlation with CD8+ T-cell presence in stromal areas (r=0.717, P<0.001), representing the strongest correlation observed in our study and indicating coordinated immune cell infiltration patterns.

### CD8+ T-Cell Distribution and Correlations

CD8+ T-cell infiltration patterns revealed significant spatial heterogeneity. Stromal CD8+ counts ranged from 0 to 96 cells per high-power field, while intratumoral CD8+ counts varied from 0 to 62 cells. The density of CD8+ T-cells was consistently higher in stromal areas compared to intratumoral regions, in line with previous reports (Ottenhof et al., 2018). This distribution pattern points to potential barriers to T-cell infiltration into tumor nests. The correlation between CD8+ T-cell infiltration and PD-L1 expression indicates an active immune response, potentially modulated by checkpoint-mediated suppression.

### Immune Marker Correlations and Spatial Relationships

Comprehensive analysis of immune marker relationships revealed complex interactions within the tumor microenvironment. PD-L1 expression in lymphocytes showed a significant correlation with FOXP3 expression in stromal lymphocytes (r=0.354, P<0.001). The spatial analysis demonstrated distinct distribution patterns, with PD-L1 expression showing heterogeneous distribution within tumor areas, while FOXP3+ and CD8+ cells demonstrated preferential localization in stromal regions. These patterns suggest organized rather than random immune cell infiltration.

### Host Response and Inflammatory Patterns

Host inflammatory response patterns showed strong associations with immune cell distributions. Cases with intense inflammation or lymphoid aggregates typically demonstrated higher numbers of both FOXP3+ and CD8+ cells in their respective compartments. This pattern indicates an organized immune response with potential implications for therapeutic targeting (Yezhong Tang et al., 2022). The presence of lymphoid aggregates often corresponded with increased immune marker expression, pointing to potential tertiary lymphoid structure formation.

These findings reveal a complex interplay between immune markers and their spatial organization within the penile cancer microenvironment. The strong correlation between stromal FOXP3+ and CD8+ cells, combined with the moderate correlation between PD-L1 and CD8+ T-cells, points to a dynamic balance between regulatory and effector immune mechanisms. These relationships have important implications for understanding immune evasion strategies and developing targeted immunotherapeutic approaches.

## DISCUSSION

Our comprehensive analysis of the immune microenvironment in penile squamous cell carcinoma reveals complex interactions between PD-L1 expression, regulatory T cells, and cytotoxic T lymphocytes. The findings provide critical insights into immune regulation within these tumors and have significant implications for therapeutic strategies.

The moderate positive correlation between PD-L1 expression and CD8+ T-cell infiltration suggests an adaptive immune resistance mechanism, where tumors upregulate PD-L1 in response to T-cell-mediated immune pressure. This finding aligns with observations in other squamous cell carcinomas (Müller et al., 2021) and supports the rationale for PD-1/PD-L1 blockade in penile cancer treatment. The variable PD-L1 expression patterns observed in our cohort indicate potential heterogeneity in response to such therapies, highlighting the need for careful patient stratification (Warli et al., 2022).

The strong correlation between stromal FOXP3+ and CD8+ T-cells represents a novel finding in penile cancer. This relationship indicates coordinated immune cell infiltration rather than random distribution, possibly reflecting an organized immune response. However, the presence of FOXP3+ cells may also indicate active immunosuppression, potentially limiting the effectiveness of CD8+ T-cell responses (Ottenhof et al., 2018). The higher density of both cell types in stromal compared to intratumoral regions suggests potential barriers to immune cell infiltration into tumor nests, a phenomenon that could impact immunotherapy efficacy.

The spatial organization of immune cells appears to be non-random and functionally significant. The preferential localization of FOXP3+ and CD8+ cells in stromal regions, combined with heterogeneous PD-L1 expression in tumor areas, points to distinct immunological compartments. This compartmentalization may influence local immune responses and could have implications for therapeutic approaches (Yezhong Tang et al., 2022). Recent clinical trials with immunotherapy in penile cancer have shown promising results, supporting the relevance of understanding these spatial relationships (Vries et al., 2023).

Our observation of varying inflammatory patterns, from minimal inflammation to intense lymphoid aggregates, correlates with immune marker expression and distribution. Cases with prominent lymphoid aggregates often showed increased immune cell infiltration, indicating potential tertiary lymphoid structure formation. These structures could serve as sites of local immune response organization and might influence treatment outcomes (Mumba, 2024). The presence of such organized lymphoid structures has been associated with improved responses to immunotherapy in other cancer types.

The weak positive correlation between tumor-associated FOXP3+ lymphocytes and tumor cell PD-L1 expression suggests a potential link between regulatory T-cell presence and immune checkpoint activation. This relationship might represent another layer of immune suppression, where multiple mechanisms work together to maintain tumor immune evasion (Ahmed et al., 2020). The complexity of these interactions underscores the need for combination therapeutic approaches.

These findings have several therapeutic implications. First, the heterogeneous expression of PD-L1 and variable immune cell infiltration patterns indicates that patient stratification will be crucial for immunotherapy success. Second, the strong correlation between stromal FOXP3+ and CD8+ cells indicates that targeting regulatory T cells might be necessary to enhance cytotoxic T-cell responses. Finally, the compartmentalized nature of immune cell distribution indicates that strategies to enhance immune cell infiltration into tumor nests might improve treatment outcomes (Stecca et al., 2021).

Study limitations include the use of tissue microarrays, which may not fully capture tumor heterogeneity, and the cross-sectional nature of the analysis, which prevents assessment of temporal dynamics in immune responses. Additionally, the varying levels of PD-L1 expression observed with different antibody clones points to the need for standardized assessment methods (Zhang et al., 2020).

Future directions should include prospective studies correlating these immune markers with treatment responses, particularly in the context of immunotherapy. Investigation of additional immune checkpoint molecules and their spatial relationships could provide further insights into immune regulation in penile cancer. The development of strategies to overcome stromal barriers and enhance immune cell infiltration into tumor nests represents an important area for future research.

In conclusion, our findings reveal a complex and structured immune landscape in penile cancer, characterized by distinct spatial organization and coordinated immune cell responses. The relationships between PD-L1, FOXP3, and CD8 expression suggest multiple layers of immune regulation that could be therapeutically targeted. Understanding these interactions is crucial for developing effective immunotherapy strategies and improving patient outcomes. The heterogeneous nature of immune marker expression emphasizes the importance of personalized approaches to immunotherapy in penile cancer.

## Data Availability

All data produced in the present study are available upon reasonable request to the authors.

